# Cross-sector Decision Landscape in Response to COVID-19: A Qualitative Analysis of North Carolina Decision-Makers

**DOI:** 10.1101/2022.03.09.22272160

**Authors:** Caitlin B. Biddell, Karl T. Johnson, Mehul D. Patel, Raymond L. Smith, Hillary K. Hecht, Julie L. Swann, Maria E. Mayorga, Kristen Hassmiller Lich

**Affiliations:** Department of Health Policy and Management, Gillings School of Global Public Health, University of North Carolina at Chapel Hill; Chapel Hill, NC; Lineberger Comprehensive Cancer Center, University of North Carolina at Chapel Hill; Chapel Hill, NC; Department of Emergency Medicine, School of Medicine, University of North Carolina at Chapel Hill; Chapel Hill, NC; Department of Engineering, East Carolina University; Greenville, NC; Department of Industrial and Systems Engineering, North Carolina State University; Raleigh, NC

**Keywords:** COVID-19, Community Health, Cross-sector collaboration, Decision-making, Crisis response

## Abstract

**Context:** The COVID-19 pandemic response has demonstrated the interconnectedness of individuals, organizations, and other entities jointly contributing to the production of community health. This response has involved stakeholders from numerous sectors who have been faced with new decisions, objectives, and constraints.

**Objective:** We aimed to examine the cross-sector organizational decision landscape that formed in response to the COVID-19 pandemic.

**Design:** We applied a systems approach to the qualitative analysis of semi-structured interviews on the cross-sector, organizational response to the COVID-19 pandemic. We analyzed transcribed interviews using conventional content analysis to synthesize key themes.

**Setting:** Semi-structured interviews were conducted via secure, video-conferencing platform between October 2020 and January 2021.

**Participants:** Forty-four state and local decision-makers representing organizations from nine sectors in North Carolina participated.

**Main Outcome Measures:** We defined the decision landscape as including decision-maker roles, key decisions, and inter-relationships involved in producing community health.

**Results:** Decision-maker roles were characterized by underlying tensions between balancing organizational mission with employee/community health and navigating organizational versus individual responsibility for reducing transmission. Key Decisions fell into several broad categories, including how to translate public health guidance into practice; when to institute, and subsequently loosen, public health restrictions; and how to address downstream social and economic impacts of public health restrictions. Lastly, given limited and changing information, as well as limited resources and expertise, the COVID-19 response required cross-sector collaboration, which was commonly coordinated by local health departments.

**Conclusions:** By documenting the local, cross-sector decision landscape that formed in response to COVID-19, we illuminate the impacts different organizations may have on information/misinformation, prevention behaviors, and, ultimately, health. Public health researchers and practitioners must understand, and work within, this complex decision landscape when responding to COVID-19 and future community health challenges.

## I. Introduction

Declared a pandemic by the World Health Organization on March 11, 2020,^1, 2^ the coronavirus disease 2019 (COVID-19) continues to rapidly spread, resulting in over 5.4 million deaths worldwide as of January 2022.^3^ COVID-19 has posed the most challenging global health crisis in at least 100 years. Specifically, the complexity of COVID-19 has been characterized by uncertain and rapidly changing information, interdependencies and feedback loops affecting decision-making across organizations and sectors, and time lags between policy changes and their ripple effects.^4,5^

Though guidance has been issued at national and state levels in the United States, the COVID-19 pandemic response has largely been carried out at the local level. Thus, to fully understand this fragmented pandemic response, it is necessary to study interconnections between local decision-makers. Due to the complexity of COVID-19, studying this local pandemic response demands a systems-approach that recognizes the distinct yet interconnected stakeholder roles shaping decisions within and across organizational boundaries.^6^ Stakeholders, defined as individuals and organizations with an interest in a given problem and its resolution,^7, 8^ range from individuals deciding whether to wear a mask to local public health officials developing and communicating guidance around mask usage.^9,10^

Recognizing that health outcomes are produced by a broad spectrum of stakeholders acting in accordance with their own goals, incentives, knowledge, and mental models of the problem at hand,^11^ we sought to study the cross-sector decision landscape emerging in response to the early- to mid-stages of the COVID-19 pandemic. We define this landscape in terms of: who is involved in decision-making affecting community health, the relationship between decision-makers’ roles and the types of decisions made, and how stakeholders influenced one other in decision-making. We conducted a qualitative analysis of organizational decision-makers in North Carolina to characterize the local decision landscape. North Carolina is a large, diverse state with several metropolitan centers. Improving health, particularly amidst crises such as this, requires coordinating complex decision landscapes. This analysis serves to illustrate a replicable approach to characterizing decision landscapes as well as to inform public health practitioners and researchers when responding to this and future infectious disease outbreaks within the context of the various perspectives, priorities, and incentives involved.

## II. Study Data and Methods

### Sample Description and Recruitment

Defining sectors as subdivisions of society including similar types of organizations serving distinct functions,^7, 12^ we interviewed state and local decision-makers from nine sectors: **business** (n=4; small business owners, real estate agent, technology company director; B1-B4), **non-profit organizations** (n=3; senior director, vice presidents (VP) of operations and risk management; NP1-NP3), **county government** (n=4; county managers, director of social services; G1-G4), **healthcare** (n=5; directors/VPs of healthcare associations, systems engineer, director of student health; H1-H5), **local public health** (n=5; local health directors; PH1-PH5), **public safety** (n=7; emergency managers, county sheriffs; PS1-PS7), **religion** (n=6; church pastors, member of church COVID taskforce; R1-R6), **education** (n=7; principal, school board member, community college president, university VP; E1-E7), **transportation** (n=3; transportation planner, traffic safety engineer; T1-T3) (**Table 1**). We defined organizational decision-makers as individuals whose job responsibilities included making decisions with a substantial impact on the organization, or individuals the organization serves.

**Table 1.**
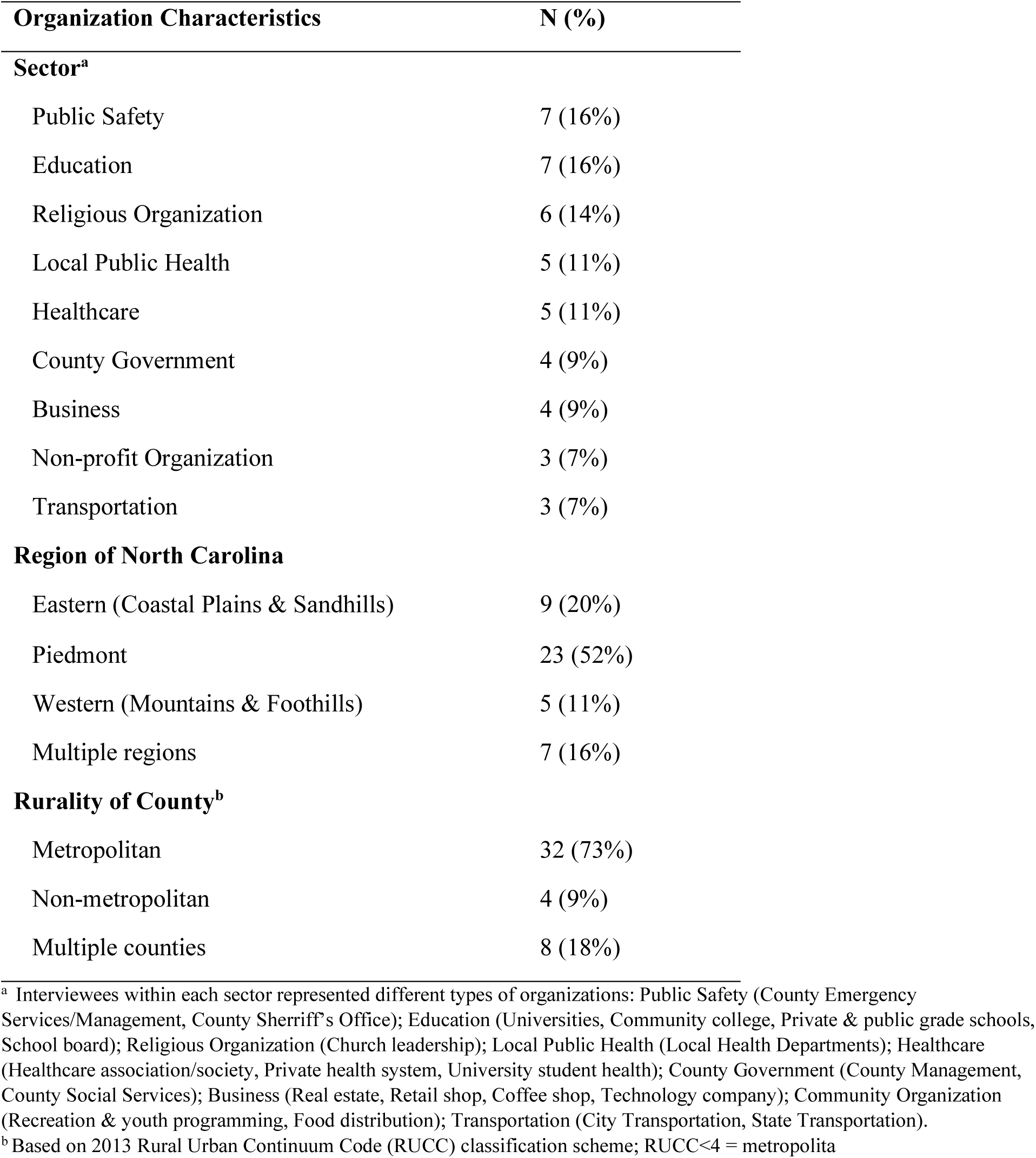
Characteristics of organizations represented in interviews with local decision-makers (N=44)

| Organization Characteristics | N (%) |
| --- | --- |
| **Sector<sup>a</sup>** |  |
| Public Safety | 7 (16%) |
| Education | 7 (16%) |
| Religious Organization | 6 (14%) |
| Local Public Health | 5 (11%) |
| Healthcare | 5 (11%) |
| County Government | 4 (9%) |
| Business | 4 (9%) |
| Non-profit Organization | 3 (7%) |
| Transportation | 3 (7%) |
| **Region of North Carolina** |  |
| Eastern (Coastal Plains & Sandhills) | 9 (20%) |
| Piedmont | 23 (52%) |
| Western (Mountains & Foothills) | 5 (11%) |
| Multiple regions | 7 (16%) |
| **Rurality of County<sup>b</sup>** |  |
| Metropolitan | 32 (73%) |
| Non-metropolitan | 4 (9%) |
| Multiple counties | 8 (18%) |

<sup>a</sup> Interviewees within each sector represented different types of organizations: Public Safety (County Emergency Services/Management, County Sheriff's Office); Education (Universities, Community college, Private & public grade schools, School board); Religious Organization (Church leadership); Local Public Health (Local Health Departments); Healthcare (Healthcare association/society, Private health system, University student health); County Government (County Management, County Social Services); Business (Real estate, Retail shop, Coffee shop, Technology company); Community Organization (Recreation & youth programming, Food distribution); Transportation (City Transportation, State Transportation).
<sup>b</sup> Based on 2013 Rural Urban Continuum Code (RUCC) classification scheme; RUCC<4 = metropolita

We used a snowball sampling approach, starting with decision-makers recommended by our research team and their cross-sector contacts, and asking interviewees for referrals to decision-makers in related organizations who may provide a meaningful and diverse perspective from their own. We interviewed 44 of the 120 potential interviewees contacted (37% response). We determined sample size by reaching thematic saturation across sectors and ensuring at least three interviews within each sector. The purpose of this sampling approach was to recruit decision-makers from diverse organizations and ensure representation across sectors and the state of North Carolina.

### Interview Procedures

Three members of the study team (KTJ, MDP, KHL) developed the semi-structured interview guide following a review of decision theory literature and iteratively revised it during the first three interviews (**Supplemental Appendix 1**). One member of the study team (KTJ), a white, male graduate research assistant with qualitative interview experience, conducted semi-structured interviews between October 2020 and January 2021 using a secure web-based video-conferencing platform. All 45-60-minute interviews were recorded and transcribed.

We asked interviewees about their perceived individual and organizational roles in the COVID-19 pandemic response. Interviewees were prompted to reflect on the key decisions that their organizations made in response to the COVID-19 pandemic early on (February and March 2020) and at the time of the interview (October 2020 through January 2021), including decisions they anticipated having to make in the near future. In discussing each key decision, we probed interviewees on the other stakeholders (within and across sectors) influencing or contributing to the decision-making process. This study was determined to be exempt from review by the UNC Institutional Review Board (#20-2087).

### Qualitative Analysis

We employed conventional content analysis to derive themes from the qualitative data.^13^ Using an inductive, iterative coding approach, we outlined a general codebook structure stemming from the semi-structured interview guide (**Supplemental Appendix 1**). We allowed interview codes and themes to emerge as two independent researchers (CBB, KTJ) coded each transcript using MAXQDA software (**Supplemental Appendix 2**).^14^ We analyzed excerpts within each code relating to the decision landscape (decision-making process codes analyzed separately), identifying major and minor themes. Decisions identified by stakeholders were coded as belonging to one or more emergent categories. Within each decision category we analyzed excerpts by sector, identifying key decision topics and documenting the interrelationships across sectors. The Consolidated Criteria for Reporting Qualitative Research (COREQ) checklist was used to guide our reporting of the qualitative analysis and results.^15^

## III. Study Results

Of the 44 stakeholders interviewed, the majority represented organizations serving constituents within a single county (primarily metropolitan), and constituencies ranged from several hundred to over 1 million (**Table 1, Supplemental Table 3**). As key informant interviewees provided organizational perspectives, individual characteristics could not be disclosed. The following themes emerged within each of three domains comprising the COVID-19 pandemic response decision landscape: (1) Perceived organizational roles, (2) Key decisions, and (3) Interrelationships between organizations (**Table 2**).

**Table 2.**
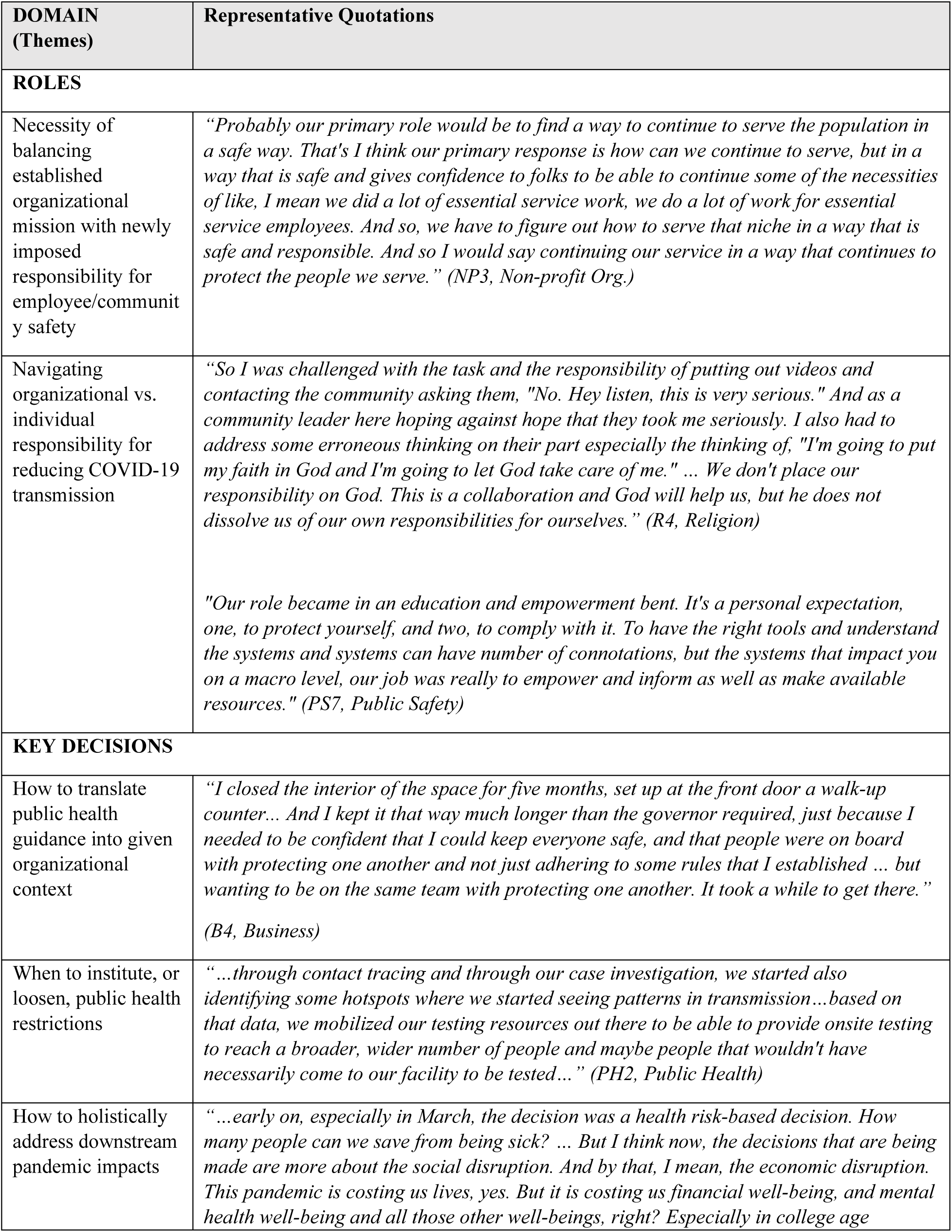
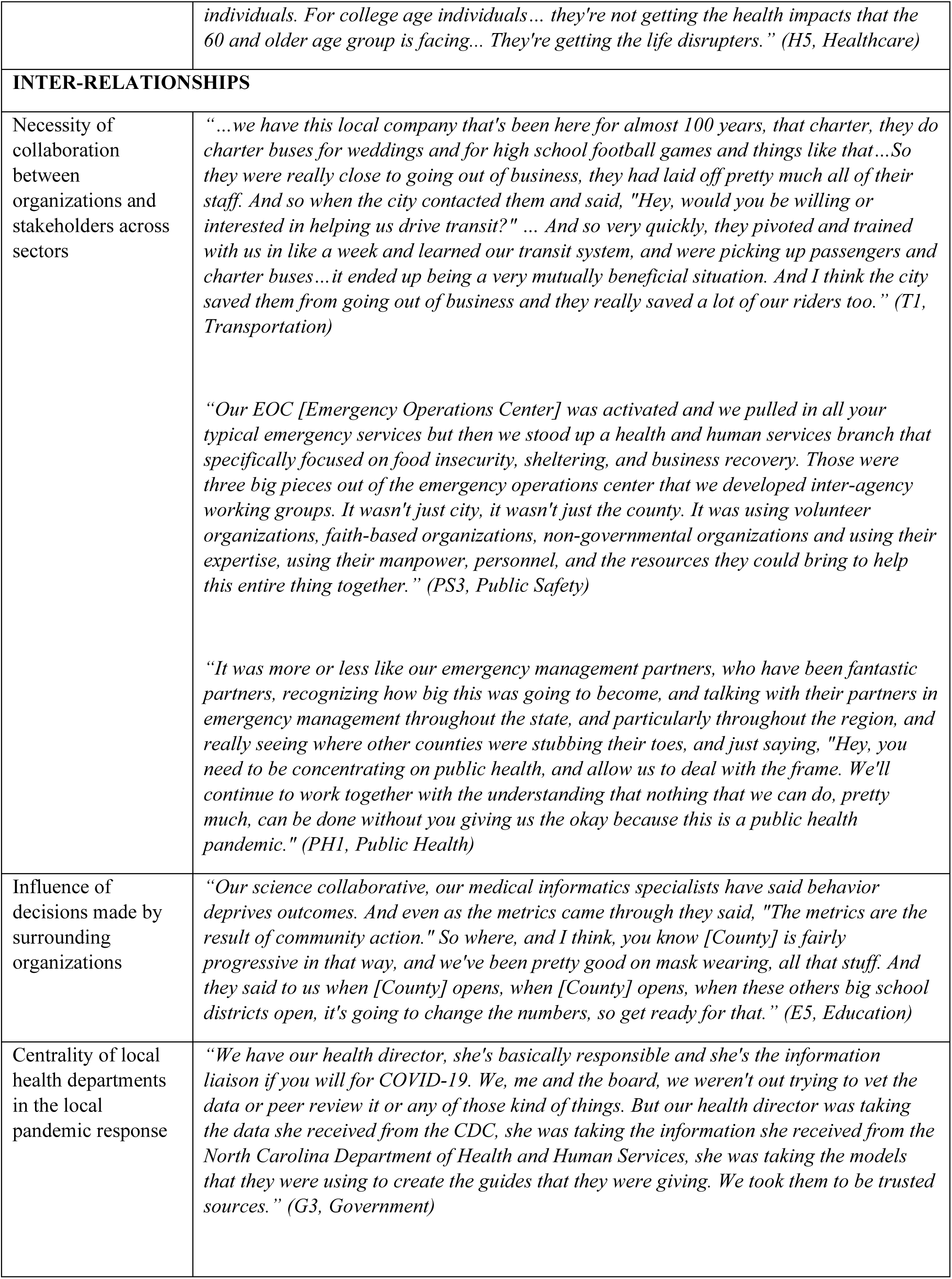
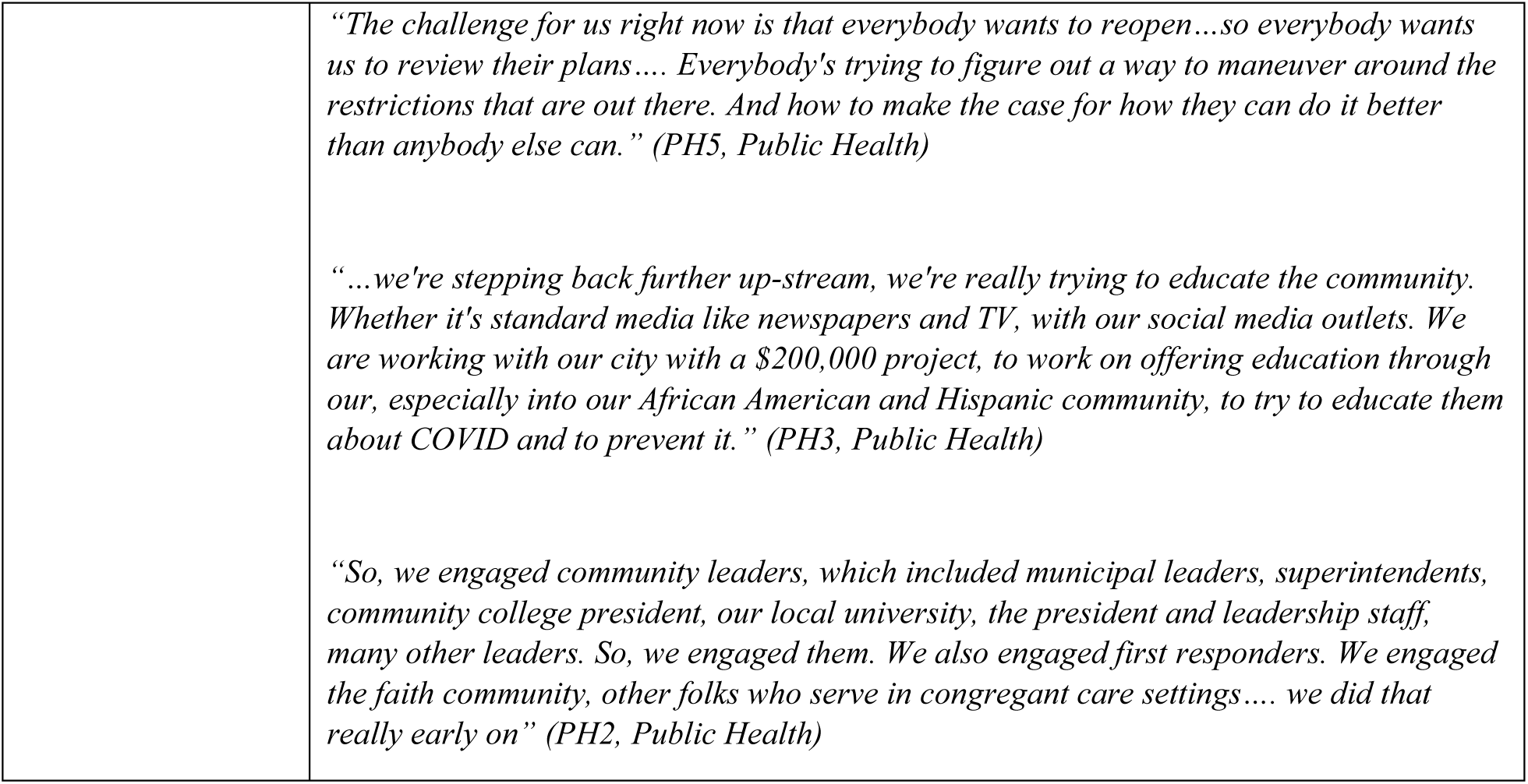
Decision landscape themes and representative quotations

| DOMAIN<br>(Themes) | Representative Quotations |
| --- | --- |
| **ROLES** |  |
| Necessity of balancing established organizational mission with newly imposed responsibility for employee/community safety | *"Probably our primary role would be to find a way to continue to serve the population in a safe way. That's I think our primary response is how can we continue to serve, but in a way that is safe and gives confidence to folks to be able to continue some of the necessities of like, I mean we did a lot of essential service work, we do a lot of work for essential service employees. And so, we have to figure out how to serve that niche in a way that is safe and responsible. And so I would say continuing our service in a way that continues to protect the people we serve." (NP3, Non-profit Org.)* |
| Navigating organizational vs. individual responsibility for reducing COVID-19 transmission | *"So I was challenged with the task and the responsibility of putting out videos and contacting the community asking them, "No. Hey listen, this is very serious." And as a community leader here hoping against hope that they took me seriously. I also had to address some erroneous thinking on their part especially the thinking of, "I'm going to put my faith in God and I'm going to let God take care of me." ... We don't place our responsibility on God. This is a collaboration and God will help us, but he does not dissolve us of our own responsibilities for ourselves." (R4, Religion)*<br>*"Our role became in an education and empowerment bent. It's a personal expectation, one, to protect yourself, and two, to comply with it. To have the right tools and understand the systems and systems can have number of connotations, but the systems that impact you on a macro level, our job was really to empower and inform as well as make available resources." (PS7, Public Safety)* |
| **KEY DECISIONS** |  |
| How to translate public health guidance into given organizational context | *"I closed the interior of the space for five months, set up at the front door a walk-up counter... And I kept it that way much longer than the governor required, just because I needed to be confident that I could keep everyone safe, and that people were on board with protecting one another and not just adhering to some rules that I established ... but wanting to be on the same team with protecting one another. It took a while to get there." (B4, Business)* |
| When to institute, or loosen, public health restrictions | *"...through contact tracing and through our case investigation, we started also identifying some hotspots where we started seeing patterns in transmission...based on that data, we mobilized our testing resources out there to be able to provide onsite testing to reach a broader, wider number of people and maybe people that wouldn't have necessarily come to our facility to be tested..." (PH2, Public Health)* |
| How to holistically address downstream pandemic impacts | *"...early on, especially in March, the decision was a health risk-based decision. How many people can we save from being sick? ... But I think now, the decisions that are being made are more about the social disruption. And by that, I mean, the economic disruption. This pandemic is costing us lives, yes. But it is costing us financial well-being, and mental health well-being and all those other well-beings, right? Especially in college age* |
|  | *individuals. For college age individuals... they're not getting the health impacts that the 60 and older age group is facing... They're getting the life disrupters.” (H5, Healthcare)* |
| **INTER-RELATIONSHIPS** |  |
| Necessity of collaboration between organizations and stakeholders across sectors | *“...we have this local company that's been here for almost 100 years, that charter, they do charter buses for weddings and for high school football games and things like that...So they were really close to going out of business, they had laid off pretty much all of their staff. And so when the city contacted them and said, "Hey, would you be willing or interested in helping us drive transit?" ... And so very quickly, they pivoted and trained with us in like a week and learned our transit system, and were picking up passengers and charter buses...it ended up being a very mutually beneficial situation. And I think the city saved them from going out of business and they really saved a lot of our riders too.” (T1, Transportation)*<br>*“Our EOC [Emergency Operations Center] was activated and we pulled in all your typical emergency services but then we stood up a health and human services branch that specifically focused on food insecurity, sheltering, and business recovery. Those were three big pieces out of the emergency operations center that we developed inter-agency working groups. It wasn't just city, it wasn't just the county. It was using volunteer organizations, faith-based organizations, non-governmental organizations and using their expertise, using their manpower, personnel, and the resources they could bring to help this entire thing together.” (PS3, Public Safety)*<br>*“It was more or less like our emergency management partners, who have been fantastic partners, recognizing how big this was going to become, and talking with their partners in emergency management throughout the state, and particularly throughout the region, and really seeing where other counties were stubbing their toes, and just saying, "Hey, you need to be concentrating on public health, and allow us to deal with the frame. We'll continue to work together with the understanding that nothing that we can do, pretty much, can be done without you giving us the okay because this is a public health pandemic.” (PH1, Public Health)* |
| Influence of decisions made by surrounding organizations | *“Our science collaborative, our medical informatics specialists have said behavior deprives outcomes. And even as the metrics came through they said, "The metrics are the result of community action." So where, and I think, you know [County] is fairly progressive in that way, and we've been pretty good on mask wearing, all that stuff. And they said to us when [County] opens, when [County] opens, when these others big school districts open, it's going to change the numbers, so get ready for that.” (E5, Education)* |
| Centrality of local health departments in the local pandemic response | *“We have our health director, she's basically responsible and she's the information liaison if you will for COVID-19. We, me and the board, we weren't out trying to vet the data or peer review it or any of those kind of things. But our health director was taking the data she received from the CDC, she was taking the information she received from the North Carolina Department of Health and Human Services, she was taking the models that they were using to create the guides that they were giving. We took them to be trusted sources.” (G3, Government)* |
| | *"The challenge for us right now is that everybody wants to reopen...so everybody wants us to review their plans.... Everybody's trying to figure out a way to maneuver around the restrictions that are out there. And how to make the case for how they can do it better than anybody else can." (PH5, Public Health)*<br>*"...we're stepping back further up-stream, we're really trying to educate the community. Whether it's standard media like newspapers and TV, with our social media outlets. We are working with our city with a \$200,000 project, to work on offering education through our, especially into our African American and Hispanic community, to try to educate them about COVID and to prevent it." (PH3, Public Health)*<br>*"So, we engaged community leaders, which included municipal leaders, superintendents, community college president, our local university, the president and leadership staff, many other leaders. So, we engaged them. We also engaged first responders. We engaged the faith community, other folks who serve in congregant care settings.... we did that really early on" (PH2, Public Health)* |

### Perceived Organizational Roles

Interviewees’ perceived roles in the COVID-19 pandemic response informed how they balanced inherent competing priorities (e.g., constituent, staff, and community safety; physical, social, and emotional wellness) in the decision-making process (**Table 3**). The following themes emerged across sectors

**Table 3.** Organization roles and key decisions among interviewees (N=44)

| **Sector (Organizations Represented)** | **Perceived Role(s)** | **Representative Decisions** |
| --- | --- | --- |
| Business<br>(Real estate, Retail shop, Coffee shop, Technology company) | Small business owners focused on continuing to meet the original mission of their business while taking responsibility for keeping customers and the local community safe. Technology company VP took on a new role within the company in the face of COVID that involved assisting external clients with COVID-related analytics. | ● Closed shop to public and built online business (Retail)<br>● Masking, distancing, and sanitizing requirements for customers and staff (All)<br>● Allowed customers to sponsor care packages to frontline workers (Coffee)<br>● Worked with governments and shipping companies on optimization (Tech) |
| Non-profit Organization<br>(Recreation & youth programming, Food distribution) | Org. leaders were tasked with managing operations and risk management throughout the pandemic. They were faced with the tension between the increased need for the services provided by their organization and the responsibility of keeping staff, volunteers and clients safe. | ● Suspended ancillary services (e.g., nutrition education) to focus on food distribution (Food)<br>● Cancelled camps and conferences (Rec)<br>● Updated volunteer safety protocols in response to changing CDC guidelines (All)<br>● Convened non-profits to support virtual learning (Rec) |
| County Government<br>(County Management, County Social Services) | Interviewees focused on ensuring the safety of their staff and direct clients. They also anticipated community needs stemming from COVID's economic impacts and worked to continue providing services (e.g., social services) in higher demand. | ● Implemented safety protocols for in-person county staff (All)<br>● Created new position to oversee food delivery for kids at home (SS)<br>● Leased new building to accommodate social distancing (Mgmt.) |
| Healthcare<br>(Healthcare association/society, Private health system, University student health) | Healthcare associations saw their role as convening organizations for: knowledge sharing, PPE allocation, and advocacy to the state. A health system systems engineer focused on the safety of providers and patients, with an emphasis on PPE allocation). The student health director saw their role as continuing to provide health care to students on campus, which expanded to include COVID testing, symptom management, and mental health care. | ● Championed stay-at-home policy in the community (Health System)<br>● Ensured continuity of care for students leaving campus (Student Health)<br>● Updated critical care resource allocation protocol (Association)<br>● Created PPE group purchasing system (Association)<br>● Supported the hiring and training of COVID contact tracers (Association) |
| Public Health<br>(Local health departments (LHDs)) | Local health directors described taking on four primary roles: (1) Communicable disease control (Testing, tracing, vaccination); (2) Supporting the translation of public health guidance into local organizational context; (3) Educating the public; (4) Convening and engaging community stakeholders to mitigate the negative impact of COVID-19 on vulnerable populations during the pandemic. | ● Issued stay-at-home order and mask mandate in advance of the state<br>● Reviewed safety protocols for local organization re-opening plans<br>● Orchestrated strike teams to address homelessness and food insecurity<br>● Worked with community partners to build vaccine champions |
| Public Safety<br>(County Emergency Services/Management) | County emergency management facilitated communication and logistics for the public health pandemic | ● Decreased number of arrests to reduce detention center volume (Sheriff) |
| , County Sheriff's Office) | response. In some cases they served as the COVID-19 incident commander, and in others they supported the LHD in this role. County sheriffs focused on ensuring the safety of their staff and people under the care of law enforcement, as well as enforcing executive orders. | ● Issued warnings for businesses not following protocol (EM)<br>● Started county fund for small business owners (EM)<br>● Oversaw logistics for mobile and community testing and contact tracing (EM)<br>● Forecasted PPE needed to run emergency operation center (EM) |
| Religious Org. (Churches) | Interviewees included church pastors, as well as one member of a church taskforce for COVID safety protocols. Church pastors described meeting the social needs of church members and the broader community, being a source of trusted leadership, taking responsibility for the safety of anyone on the church property, continuing to instill hope in community, and overseeing the church's financial situation. | ● Suspended (and in some cases, later resumed) in-person religious services<br>● Assigned ushers to control the flow of people to maintain social distancing in in-person services<br>● Identified gaps in community social services and worked with other groups to meet those needs<br>● Partnered with LHD to host testing event<br>● Installed new air purification systems |
| Education (Universities, Community college, Private & public grade schools, School board) | Despite interviewees holding different roles, they fairly universally described their role as promoting the well-being of students through continuing education (in varying forms), meeting social needs of students' families and surrounding communities, and ensuring student safety. | ● Transitioned to remote learning (All)<br>● Hired COVID coordinators at each school responsible for temperature and symptom checks (Primary, Secondary)<br>● Delivered laptops and hotspots to students (Primary, Secondary)<br>● Developed testing and isolation procedures for bringing students back (Post-secondary) |
| Transportation (City Transportation, State DOT) | Interviewees in city and state transportation saw their role as ensuring safety of citizens while using public transit, public spaces, and roadways. | ● Transitioned public input sessions to be virtual (All)<br>● Hired private transportation company to supplement/avoid cutting routes (City)<br>● Lent businesses public space for outdoor dining (City) |

#### Necessity of balancing established organizational mission with newly imposed responsibility for employee/community safety

Interviewees from all sectors prioritized customer, constituent, and community safety, often as a new responsibility in addition to their originally stated missions. For example, an interviewee from a non-profit dedicated to youth and recreational programming emphasized the challenge of carrying out this mission when they could no longer bring the community together in-person. In this case, the organizational mission and the responsibility for community safety were viewed as being in tension with one another; however, other interviewees viewed keeping their constituents safe as consistent with their original organizational mission, which became *“more urgent than ever before”* (R1, Religion). This responsibility also extended to the health of the broader community. *“The safer we are here, the safer folks are in the community”* (R2, Religion).

#### Navigating organizational vs. individual responsibility for reducing COVID-19 transmission

Given that many COVID-19 safety protocols required individual behavior change, interviewees acknowledged the limitations of their organizational roles in enforcing these measures. However, they underscored their role as being to educate and empower the public to uphold their personal responsibilities in mitigating COVID-19 spread. “*It’s a personal expectation, one, to protect yourself, and two, to comply with it*…*Our job was really to empower and inform as well as make available resources*” (PS7, Public Safety). One pastor disseminated educational videos to combat misinformation – “*This is a collaboration and God will help us, but he does not dissolve us of our own responsibilities for ourselves*” (R4, Religion). The form of education varied and was often tailored to communities. Interviewees emphasized the importance of ensuring that constituents understood why public health measures were needed. Empowerment included leadership modeling public health behaviors and securing the resources, such as masks, to support community health-minded decisions.

### Key Decisions

Fulfilling the roles described above involved decisions related to continuing or suspending in-person services, instituting safety protocols, allocating resources (human and physical), testing/screening, contact tracing, and vaccination. Interviewees described a decision ecosystem in which the consequences of one decision (whether related to viral transmission, economic impacts, or organizational realities) prompted the need for subsequent decisions. A full matrix of COVID-19-related decisions described is included in **Supplemental Appendix 4** and summarized in **Table 3**. The following thematic decision categories emerged

#### How to translate public health guidance into organizational context

All interviewees made decisions to discontinue, or transition remotely, all non-essential in-person services in March 2020, informed by state and local stay-at-home orders. Though this was framed as a necessarily cautious response to the uncertainties of the pandemic, it prompted a cascade of decisions related to translating guidance into organizational contexts to maintain services/mission while ensuring employee and community safety. Decisions included distinguishing essential vs. non-essential personnel to inform remote work scheduling, securing PPE for essential personnel, and securing the technology necessary to support remote work. Even Local Health Directors (LHDs) had to make internal staffing and protocol decisions, all while being propelled into a more central role than ever before. “*A big part of my workforce have children… How do we work and show up to serve the community while balancing the needs of what you’re having to do at home?*” (PH4, Public Health).

In contrast, re-opening decisions were more contentious. While many strove to re-open, some decision-makers remained closed or instituted safety protocols beyond legal mandates. “*I needed to be confident that I could keep everyone safe, and that people were on board with protecting one another*” (B4, Business). However, pressure from community members to re-open grew over time. “*I’ve watched some of my colleagues at more conservative schools have to make decisions that they weren’t 100% comfortable with, in terms of how rooms were organized, in terms of mask use …because of the pressure of their community*.” (E3, Education).

#### When to institute, or loosen, public health restrictions

While not all sectors were directly involved in testing, tracing, and vaccination, related decisions made by LHDs and Emergency Managers (EMs) influenced community transmission, and thus decisions about re-opening and safety protocols by organizations in other sectors. LHDs and EMs instituted contact tracing early on. “*To date, we believe that we maintained a seven-day rolling average of less than a hundred cases a day because we continue to do contact tracing*.” (PS4, Public Safety). LHDs and EM also implemented testing, often in partnership with external clinical entities; however, interviewees described challenges in carrying out these services equitably at scale. “*Contact tracing in most public health agencies wasn’t fit for purpose, for the scale*.” (B2, Business)

#### How to holistically address downstream pandemic impacts

A final category of decisions related to developing new or extending existing services to address social impacts of COVID-19 restrictions, such as homelessness and food insecurity. In some cases, this meant balancing infection risk with health risks of downstream consequences. Interviewees noted a primary tension in that efforts to “*dampen down COVID in our community are also the things that are putting some of our most vulnerable population at risk*” (PH5, Public Health). For organizations working to meet social needs, the recognition of heightened need motivated organization leaders to ensure services continued, even if processes had to change to keep staff, volunteers, and constituents safe. “*There’s a whole litany of things that have kept us busier and have really proven the urgency and the significance of community-based and faith-based organizations*.” (R1, Religion).

### Interrelationships

The complexity and novelty of COVID-19 demanded the pooling of resources and expertise in decision-making, exemplifying the interrelationships between individuals, organizations, and resources within and across sectors.

#### Necessity of collaboration between organizations and stakeholders across sectors

Interviewees described creatively responding to COVID-19-imposed challenges by forming new and developing existing collaborations, and bringing together diverse stakeholder perspectives, to prevent blind spots in decision-making. Three main categories of collaborations were identified: (1) **Public – Public**, particularly partnerships within sectors of local government (e.g., public health and EM co-leading the local pandemic response), (2) **Public – Private**, particularly government-initiated partnerships with non-governmental organizations (e.g., county social services partnering with community organizations to distribute COVID federal relief funds), and (3) **Private – Private**, particularly among businesses, non-profits, and religious organizations (e.g., local businesses partnering to deliver care packages to frontline workers). Interviewees universally described feeling that their collaborative capacity became stronger as a result of COVID-19, “*One of the positives that’s going to come out of COVID is that we’re going to have a more robust, cohesive, collaborative model of nonprofits and organizations working together*” (R1, Religion).

Interviewees also described the impact of decisions made by the public and other surrounding organizations. As one interviewee noted in reference to the influence of community mask compliance and school district re-openings, *“metrics are the result of community action… If we change our behavior, it’s going to change the numbers”* (E5, Education). In addition to influencing COVID-19 transmission trends, local decisions were described as influencing the feasibility of asking employees, volunteers, or customers to return in-person (e.g., Are schools open to provide childcare? Is public transportation running at full capacity?).

#### Centrality of local health departments in the local pandemic response

Central to many of the interrelationships described by interviewees, LHDs served a critical function in the pandemic response, both informing local decision-making and facilitating the implementation of higher-level decisions through collaboration with other sectors. LHDs served four primary roles, each of which involved decision-making: (1) Directly responding to the communicable disease outbreak (e.g., testing, tracing, vaccination); (2) Guiding the translation of public health guidance into local organizational contexts; (3) Educating the public; (4) Convening and engaging community stakeholders (**Figure 1)**. Implementing a comprehensive pandemic response required collaborating with other sectors, such as hosting testing and vaccination events in parking lots. LHDs informed decisions at the crossroads between federal- and state-level guidance and local organizations. They were viewed as “trusted sources” (G3, Government), providing tailored public health advice, visiting local businesses, and reviewing safety protocols. Educating the public required monitoring and reporting local COVID-19 trends through data dashboards and collaborating with leaders from other sectors to host press conferences and conduct educational campaigns. Lastly, LHDs were tasked with convening and connecting stakeholders across sectors to ensure the inclusion of diverse perspectives in addressing the economic and social determinants of health, creating *“better health through better partnerships”* (PH3, Public Health).

**Figure 1.**
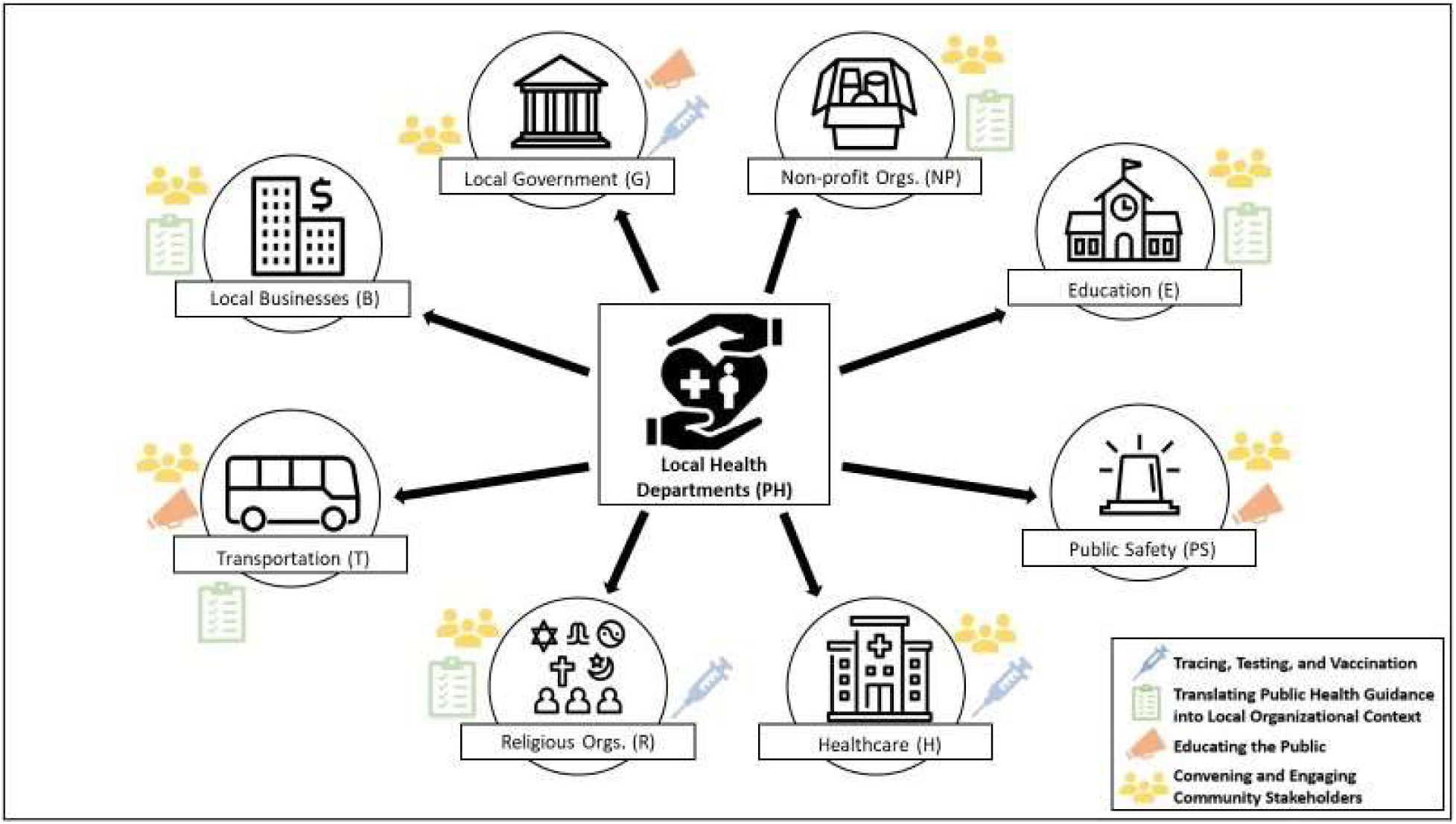
Central roles of local health departments in coordinating local COVID-19 pandemic response across sectors Local health departments were central to the local pandemic response. This figure shows how they informed and facilitated decision-making related to each of four roles in collaboration with other sectors.

## IV. Discussion

The COVID-19 pandemic has thrust decision-makers across sectors into new roles in a public health crisis response, creating a decision landscape with numerous actors and varying levels of coordination between them. In response to the complexity of COVID-19, decision-makers engaged in both collaborative and semi-autonomous decision-making processes and depended upon new authorities, especially LHDs. In this resulting “polycentric” decision-making system, public and private actors worked at different scales to collectively produce a pandemic response.^16^ By outlining the local decision landscape in the COVID-19 pandemic response, this study informs public health researchers, practitioners, and organizational decision-makers in how to navigate this and future complex, cross-sector population health challenges.

This study builds off prior work highlighting cross-sector responses to crises such as Hurricane Katrina and H1N1.^17, 18^ However, this study is the first, to our knowledge, to investigate such a broad, cross-sector decision landscape in response to COVID-19. Several prior studies have investigated decision-making in response to COVID-19 within single sectors. These studies support the decision categories that emerged from our analysis, including decisions related to allocating resources,^19^ translating guidance into real-world organizational context,^20^ and addressing downstream social impacts.^21^ Our finding that cross-sector collaborations were critical components of the COVID-19 pandemic response builds upon several prior studies illustrating specific collaborations emerging in response to COVID-19-related needs, ranging from childcare for healthcare workers to local COVID-19 surveillance through school districts.^22-25^

In line with our findings, prior work has emphasized the importance of community engagement in comprehensive pandemic responses and the necessity of communicating with stakeholders amidst changing, uncertain information.^26^ Challenges with community-based approaches, however, include balancing the need to respond quickly with the time it takes to meaningfully garner stakeholder perspectives.^27^ The need to navigate complex tradeoffs and often conflicting priorities within a community further underscores the importance of a cross-system governance or organizing structure with input from many stakeholder groups.^21^ Given the need to act quickly, communities should agree on such structures in advance of public health crises. Our analysis highlighted the importance of LHDs serving as what “Public Health 3.0” defines as a “chief health strategist”,^28^ working with other organizations directly and indirectly to govern the local public health system.^29^

The decision landscape emerging in response to COVID-19 has implications for efforts to promote population health, beyond the immediate context of COVID-19. Though a global pandemic uniquely affects all individuals and organizations, other population health challenges operate within complex systems, influenced by multi-level determinants, ranging from individual action to social policy.^30^ This can create inconsistent priorities and decisions within communities that block progress. The role of stakeholders across sectors in the pandemic response, and the interrelationships between these sectors, support the growing call for the importance of cross-sector collaboration in promoting population health.^7, 31, 32^ Our findings further align with the vision of “Public Health 3.0” to expand the reach and scope of public health to “address all factors that promote health and well-being, including those related to economic development, education, transportation, food, environment and housing.”^28^ Public health leaders advocating for this broadened definition of public health have underscored that carrying out this vision successfully requires the development of sustainable cross-sector partnerships, community engagement, and the application of a systems perspective to problem solving.^33^

The “10 Essential Public Health Services” also reflect this reality, which considers the public health system to include not only public health agencies and healthcare providers, but also public safety, human services, and education, among other sectors.^34^ The decisions described in our analysis broadly fall into the three core functions: assessment (e.g., contact tracing, testing), policy development (e.g., implementation of executive orders, mobilizing community partnerships, educating the public to support effective policy change), and assurance (e.g., workforce maintenance, ensuring equitable access to services).^35^ However, the COVID-19 pandemic has showcased that the centrality of equity in the revised essential services may still be aspirational. Disparities in COVID-19 morbidity and mortality rates by race and socioeconomic status underscore the need for system-wide decision-making that better prioritizes equitable access to health services, ranging from healthy living conditions to clinical care.^36, 37^ Additionally, the pandemic has highlighted the importance of the essential service, to “build and maintain a strong organizational infrastructure for public health,” moving forward. Bringing together the many sectors involved in the United States’ fragmented public health system effectively and sustainably, beyond the immediate aftermath of a crisis, requires local foundational infrastructure supporting timely and comprehensive data collection;^38^ flexible funding mechanisms that recognize the necessity of cross-sector work in public health;^39^ and sufficient staffing capacity, particularly in response to the burnout of the current public health workforce.^40, 41^

These findings should be viewed in the context of several limitations. While we were intentional in ensuring diverse representation of interviewees across sectors, organization type, and geography (across North Carolina), the sample does not represent an exhaustive list of organization types involved in the COVID-19 response. The snowball sampling technique employed increases the potential that the opinions uncovered were more homogenous than they would be otherwise. However, we were explicit when asking for recommendations that we were interested in uncovering a more complete and broader perspective on the subject. Thematic saturation was based on generalizable themes that emerged across sectors. Future research should investigate specific instances of cross-sector collaboration, interviewing more stakeholders involved, to gain a more detailed understanding.

The timing of interviews with respect to official guidance, transmission rates, and vaccination rollout undoubtedly influenced participant responses. We incorporated timing into interviews and analysis. Additionally, participant responses may be subject to self-report bias, given limitations of recall and the potential for selective reporting. Lastly, decision-makers willing to participate in public health research may have differed from those who refused in the extent to which they valued and trusted scientific information. However, participants described a range of perspectives on how they incorporated scientific information into decision-making.

This analysis of local decision-makers from nine different sectors in North Carolina documents the complex, cross-sector local decision landscape in response to the COVID-19 pandemic. Most notably, this analysis highlights the expanded roles of decision-makers across sectors in the pandemic response, the key types of decisions faced, and how decision-makers relied on collaboration and the guidance of LHDs. Understanding this decision landscape serves to inform public health researchers and practitioners about who is involved in decision-making related to community health and how. Knowing this can support communities in collaborating to improve organizational decision-making processes with community and population health in mind. It also underscores the need for public health infrastructure to improve information dissemination, priority setting, and alignment in response to future crises and other complex health challenges.

## V. Implications for Policy & Practice

- The COVID-19 pandemic response has involved decision-making by organization leaders from across sectors (e.g., business, government, non-profit organizations, public health), all of whom contributed to community health in inter-connected but not fully coordinated ways.
- By influencing local decision-making across diverse sectors and facilitating the implementation of higher-level decisions through collaboration, local health departments executed chief health strategist responsibilities. Building local health department capacity for this work is critical to the success of future crisis response.
- Given the reality that community health, during crisis response and otherwise, is influenced by numerous sectors, public health must develop an infrastructure to facilitate cross-sector coordination. This includes the ability to communicate public health priorities and seek value alignment in ways that respond to the diverse needs and levels of understanding across sectors.

## Data Availability

Data produced in the present study are available upon reasonable request to the authors.

## Acknowledgements

The authors are grateful to all of the individuals who participated in the interviews for this analysis. They are also grateful to Paul Mihas, who provided formative qualitative research guidance.

## Supplemental Materials

**Supplemental Appendix 1**. Semi-structured Interview Guide

**Supplemental Appendix 2**. Final codebook used in conventional content analysis

**Supplemental Appendix 3**. Interviewee characteristics

**Supplemental Appendix 4**. Spectrum of COVID-19-related decisions faced by local North Carolina decision-makers across sectors

