## Supplemental Appendices for "Cross-sector Decision Landscape in Response to COVID-19: A Qualitative Analysis of North Carolina Decision-Makers"

**Supplemental Appendix 1.** Semi-structured Interview Guide

**Pre-COVID-19**

*Intro:* *We’d first like to ask a few general questions about the background of your organization, without yet considering the COVID-19 pandemic.*

- Could you please describe, in your opinion, your organization’s mission?
  - What main services do you provide the community?
  - Which communities do you serve?
- What is your role in the organization?
  - What are your primary responsibilities?
  - Who are the people you report to?
  - Who are the people who report to you?

**General COVID-19**

*Let’s now transition to the rest of our questions, those which deal specifically with the COVID-19 Pandemic. We’d first like to ask a few questions about how your organization approached the Pandemic.*

- How does your organization talk about the risks from COVID-19?
  - Risks regarding the potential spread of infection within your organization
  - Risks regarding the potential spread of infection from people in your organization to those your organization interacts with.
- Can you describe how you see your organization’s role regarding the pandemic during?
  - Can you describe your organization’s role regarding reducing risk from the pandemic?
    - Within the organization?
    - Within the wider community?
  - How do your organization see its role in developing safety protocols?
    - Responsibility?
  - How does your organization see its role in ensuring testing and tracing?
- Can you describe the thoughts you have on people’s individual responsibility to reduce risk?
  - How do the responsibilities of your organization interface with people’s individual responsibility to reduce risk?
  - How do your personal perspectives on risk and risk reduction compare to your organization’s perspectives?
- Are there ways in which you believe your personal perspective on risk and risk reduction differs from the perspective of your organization?

**Early (March-April) COVID-19**

*Next, we’d like to ask a few questions about decisions and decision-making in your organization during the context of COVID-19.*

*We’d like for you to walk us through some specific decisions your organization has had to make during the COVID-19 pandemic and how those decisions were made.*

- In your organization, what key macro decisions did you make as an initial response to COVID-19 back in March-April? (Please consider decisions that had a big impact on the organization or those whom the organization effects.)
  - Safety measures?
  - Staffing changes?
  - New services?
  - Opening-up / closing-down protocols?
  - Anything regarding social distancing, mask usage, public relations messaging
- Can you walk me through how (one of these key decisions) were made?
- What alternatives were you choosing among?
  - What stakeholder considerations were you weighing when deciding between options?
  - What were the most important outcomes that are informing the decision?
  - How certain were you of the future outcome of the decision? How have you sought to gain more certainty?
  - What parties/stakeholders were involved in those decisions?
  - What evidence/information/data was used to make the decision?
- Do you wish you could have formalized the decision-making process during that time?
- What was the most difficult thing about organizational decision-making during the first few months of the Pandemic? How could these difficulties be overcome, or how were they overcome for your organization?

**Current/Future COVID-19**

*Now we’d like to shift our focus to the current and future context of the Pandemic.*

- How have discussions regarding your organization’s role changed since last spring?
  - How has your org’s perception of risk changed since last spring?
- How is your organization talking about what might happen in the next several months?
- In your organization, what key macro decisions are you currently considering, say over the next 3-6 months? (Please consider decisions that had a big impact on the organization or those whom the organization effects.)
- Can you walk me through how (one of these key decisions) is being made?
  - What happened first?
  - Who was involved?
  - What happened next?
  - What, if anything, do you wish had been different about that decision?
  - What alternatives are you choosing among?
  - What stakeholder considerations are you weighing when deciding between options?
    - Public interest
    - Public opinion
    - Investors/funders
    - Bad/good press
    - Customer/consumers/participants
    - Organization staff
  - What are the most important outcomes that are informing the decision?
  - How certain are you of the future outcome of the decision? How are you seeking to gain more certainty?
  - What parties/stakeholders are involved in those decisions?
    - What unique roles do each contribute to the decision-making process?
  - What evidence/information/data is being used to make the decision?
    - What is your greatest need for evidence/information/data?
    - What is the best way in which information/data or other inputs could be presented to inform this decision?
    - Do you think it’s the responsibility of the organization or a separate authority to provide this information?
- What is the most difficult thing about organizational decision-making right now? How could these difficulties be overcome, or how are they being overcome for your organization?
  - Too much information to process?
  - Information keeps changing?
  - Unclear who the proper authorities are?
- Do you wish you could have formalized the decision-making process?
  - Can you tell me more about that?

*Finally, we’d like to discuss how the decision-making process in your organization may have changed during COVID-19.*

- Before COVID-19, what was the general decision making process like?
  - How were decisions made?
  - How hierarchical was the decision-making process?
  - What was a typical timeline for decision-making?
  - Who made the final decision?
- How has the general decision-making process changed during the COVID-19 pandemic, if at all?
  - More hierarchical? New staff roles?
  - Task forces developed?
  - Faster pace than normal?
- How have these inputs/factors you consider in the decision-making process changed during the COVID-19 pandemic, if at all?
  - New authorities? New inputs?
  - New staff dedicated to processing inputs?
  - Have these inputs changed during the course of the pandemic?

*Closing: We greatly appreciate your input… We’re aiming to learn from a wide range of perspectives and collect a diversity of opinions.*

- Thinking back on your answers, is there anyone else in your organization or related organizations you know of whom we should talk to get a more complete picture or a differing perspective on this topic?

**Supplemental Appendix 2.** Final codebook used in conventional content analysis (* indicates codes included in this paper)

| **Domains & Codes** | **Description** |
| --- | --- |
| **Domain 1:**  **Organization Background** | These codes should describe the organization in which the interviewee works and makes decisions. The goal with these codes is to capture the organizational features which both present particular decisions to the individual and frame the process of decision-making. These codes will also capture how the organization understands itself with respect to its role and responsibility in responding to COVID-19. |
| Individual's role in Organization* | What capability does the interviewee have to make decisions in the organizations; who responds to them in the decision-making process and who do they report to; what kinds of decisions the individual is responsible for making, either alone or with the support of others in the organization. |
| Organization’s COVID-19 role* | What niche the organization sees itself occupying in the COVID-19 response, whether as pertains to its role in the community (external) or role for the organization itself (internal). Includes organization's perceived responsibility (how the organization views individual and collective responsibility during COVID-19. This responsibility can be considered with respect to, for instance, the need to adhere to safety protocols or ensure one’s personal level comfort with social distancing and mask usage.) |
| Individual's COVID-19 role* | Interviewee's role in the organization's COVID-19 response |
| Organization services* | Characteristics of the organization that are related to what the organization does or provides. Include overall mission with this code |
| Organization staffing* | Who composes the organization |
| Organization constituents* | Who the organization works for or serves |
| **Domain 2:**  **Decision-making Context** | These codes should describe the context—political, epidemiological, social/communal—in which decisions were being made. Coding these items will serve to outline the backdrop against which alternatives were being considered. These items may also serve to further understand the constraints decision-makers were forced to deal with. |
| National politics | Any commentary on topics related to national politics, whether or not directly related to any particular decision. Include commentary on social unrest more broadly with this code. |
| State politics | Any commentary on topics related to state politics, whether or not directly related to any particular decision. |
| County politics | Any commentary on topics related to local/county politics, whether or not directly related to any particular decision. |
| Pandemic trends/evolution | How the pandemic was progressing during the moment of the decision—what were trends like in their own or other counties, states, and countries? These trends may correspond to those specific to the virus (e.g. what wave was being experienced, where incidence levels increasing or decreasing) or those specific to the healthcare system (e.g. surge capacity, overwhelming number of hospitalizations, lack of treatment facilities). |
| Landscape of Other Organizations* | What other major organizations exists within sufficient proximity to the main organization of interest that could affect decision-making. Include in with this code any key decisions made by other organizations. This should only be used when there is no partnership between the orgs being discussed and when they are different types of organizations. |
| Challenges | If identified as such, the most challenging components of the decision-making context (e.g. the timing of decisions, mix of information). This code will likely overlap with other codes within this section, or even codes in other sections (e.g. when a challenge is process-related) |
| Community Opinion | What were conversations like within the community that the organizations inhabit or serve? How were people talking about the pandemic, what were their behaviors like and assumptions they had about its trajectory? How was social media/news media within the community contributing to the decision-making context, if at all? How did community members relate to the authority of decisions being made? |
| Legal Constraints | Legal obligations the organization has to follow which inform or constrain the decision-making follow (either general laws or those specific to the organization as such) |
| Information Quality/Certainty | Commentary on the credibility of information being used to inform decisions and the pace at which new information (and potentially conflicting or changing information) is coming out. Or commentary on the level of certainty in the information being used to inform decisions |
| **Domain 3:**  **COVID-19-related Decisions** | These codes should capture the “landscape of decisions” being considered during COVID-19. More detailed dimensions of these decisions will be considered in a separate section. |
| Safety protocols* | [When people were in person] Was the organization deciding whether or not to adopt specific safety protocols for individuals within their organization or individuals their organization serves? Or were they providing recommendations for safety protocols to other organizations? These protocols may include but are not limited to those pertaining to masks usage, social distancing, internal ventilation systems, interior design (e.g. installation of plexiglass), etc. |
| Population served/services* | Was there something about the pandemic that prompted the organization to consider new services it would offer (either it’s own employees and staff or those whom it serves) or new populations that would now receive typical services? This may include the development of new facilities to meet COVID-19 demands or even the prioritization of a particular population to receive a particular set of services. These items would be an extension or greater focus on the kind of services typically delivered or the kinds of populations typically served by the organization. |
| Population served/services 🡪 Underserved populations* | When commentary on services or populations serves applies to underserved or marginalized communities, including all community of ethnic minorities. Make sure to only apply this when it is related to a decision being made |
| Testing/Tracing* | Any decision made about the expansion or implementation of testing or tracing strategies |
| Vaccine* | Anything pertaining to a decision the organization needed or needs to make about the vaccine—including but not limited to which individuals would receive the vaccine, how details about the vaccine would be communicated, how the vaccine would be delivered, etc. |
| Continuing/Suspending In-Person Services* | Any decision strictly about the opening or closing of a building for the sake of avoiding in-person contact. This may pertain to the opening or closing of the organization itself, or ways in which the organization provided advice to other organizations about whether to open or close. The decision itself may also not be a binary between opening and closing, but rather what technologies to establish to maintain services even while individuals could not meet in person due to closures (e.g. transitions to partial zoom gatherings, delivery of services through digital means as opposed to in person, etc.) |
| Physical resource allocation* | Given changes that occurred during COVID-19, any decision pertaining to the new allocation of physical resources (e.g. equipment, monetary resources) to adapt to those changes. The allocation of resources may be considered either internal to the organization (e.g. the allocation of masks or safety equipment to personnel) or with respect to the organization’s constituents (e.g. the allocation of medical equipment to different hospital systems). Codes such items even if they are not mentioned explicitly but if new service requires allocation of resources |
| Human resource allocation* | Given changes that occurred during COVID-19, any decision pertaining to the new allocation of staff/workforce to adapt to those changes. This may include re-allocating existing workforce to new/different tasks or hiring additional workforce for additional needs arising as a result of COVID. |
| **Domain 4:**  **Decision-Making Process** | These codes should capture the dynamics of the decision-making process itself, including different moments in the evolution of the decision, the form by which the decision was made, and ultimately the outcome of the decision and how that decision was evaluated. |
| Mutual Partnership* | Any commentary about an organizational partnership in which both organizations benefit. Apply liberally, even if both partnerships are not involved in decision-making |
| Dependent Partnership: LHD* | Trumps other dependent partnership codes if discussing working with or relying on guidance from LHD |
| (Non-LHD) Dependent Partnership: Information Sharing* | Any commentary about an organizational partnership in which one organization only benefits from the information provided by another organization |
| (Non-LHD) Dependent Partnership: All other resources* | Any commentary about an organizational partnership in which one organization only benefits from the services provided by another organization |
| Communication between decision-makers* | Explicit or implicit commentary on the flow of communication between parties involved in the decision-making process. Include both communication between decision-makers across similar orgs and communication between decision-makers within single org |
| Decision conflict | Were there explicit moments in which conflict arose in the decision-making process? Whether conflict internal to the organization or between two or more organizations? Similarly, were there moments in which conflict was explicitly resolved? Anything related to the details of the conflict itself or the process by which it was resolved should be included in this code. |
| Decision resolution | When working with multiple different organization, multiple offices within the same organization or multiple types of information, how was consensus reached on a decision characterized by disagreement? How were the perspectives of multiple different actors and multiple different types of information and evidence considered and aggregated, if at all? |
| Pace of Decision-Making | How did the decision-maker or organization consider the pace with which decisions had to be made? What variables informed this pace? How did this pace informed the kinds and qualities of decisions that were made or are being made? |
| Learning | Any explicit description of how the organization learned and improved upon its decision-making process throughout the pandemic |
| Structure Formality | If formally described, what was the structure of the decision-making process? Was it described as more systematic or chaotic? Were there unique groups developed to facilitate the decision-making process (e.g. task forces)? Was the decision-making process informed by any set of rules or follow a specific learning cycle? In general, was the process hierarchical or diffuse? |
| **Domain 5:**  **Decision Inputs** | These codes should capture what kind of information was most influential in the decision-making process and what kind of information was explicitly not considered during the decision-making process. They also capture what were the beliefs or assumptions about particular kinds of information. |
| Stakeholders (not at the table) | Who were the primary stakeholders that were considered during the decision-making process but not involved in making the actual decision? Internally, this may include staff and employees, boards/committees, and constituents/costumers/consumers. Externally, this may include public opinion (including professional journalists and critiques) and stockholders. This code should also cover the assumptions the organization had about stakeholder beliefs/opinions. |
| Stakeholders (at the table) | What stakeholders were involved in making the decision? These should primarily be the decision-makers |
| Missing/Desired information | Were there particular pieces of information that the organization desired during the decision-making process but was ultimately not available? |
| Official Guidelines | How did official guidelines (whether drafted by government agencies or other authoritative voices) influence the decision-making process? Which guidelines were considered? These guidelines may include, for instance, those prepared by county, state and federal officials. |
| Data | Any commentary on data that was used or tracked by the organization to inform decision-making. Include instances in which there are commentaries (positive or negative) on the general use of data in decision-making |
| Scientific authority/expertise | What kind of scientific authority informed the decision-making process and how? This could include scientific publications (e.g. what expert scientists publish – deference to the science itself) or scientific expertise (e.g. what expert scientists say – deference to the person). This code is distinct from whatever science ultimately incorporated itself into official guidelines, which should be coded in the “official guidelines” code. This code should also include the beliefs and assumptions the organization had about scientific knowledge, including specific items of knowledge such as the risk of disease spread, the risk of morbidity and mortality, and the expected utility of medical interventions such as vaccines and antiviral treatments. This code should not include anything related to modeling (see next code). This should pertain to either specific conversations interviewees had with scientists or specific scientific publications (i.e. NOT general guidelines). |
| Past experience | Refers to experience prior to COVID that may inform how decisions during COVID are approached |
| Financial considerations | Refers to consideration of the financial/budget impact of various decisions on the organization |
| Modeling | Any input related to modeling work, including the assumptions and beliefs they have about modeling as a scientific practice. |
| Peer decisions | How did the decisions made by peer institutions (e.g. institutions in nearby counties, states, etc.) influence the decisions made by the organization in question? How much dependence was there upon peer decisions? |
| Values | What values of the organization strongly informed the decision-making process? This could include items such as financial stability, concerns for equity/justice, concerns for mental health and safety, the well being of the community or the organization’s constituents, concerns for spirituality, and political alignment with broader political movements. Interviewees don't need to explicitly state these priorities as values, so long as their comments reflect latent values within the decision-making process. |
| **Domain 6:**  **Post-Decision Reflections** | These codes should capture reflections on the process following a decision being made, including how that decision was communicated to those affected, how the outcomes of that decision were perceived, and how lessons learned were incorporated into future decisions. |
| Communication/Education* | How were decisions or the decision-making process communicated by the organization? This communication could be either to folks within the organization (internal) or to folks outside the organization (external) such through offices of public relationships or through social media. How did the organization understand the importance of communication for disseminating information about decisions after they were made? |
| Outcomes (success/failure/regret)* | How were the outcomes of decisions ultimately considered for the decision maker? Were there particular decisions that were considered failures or successes? Were there decisions that they regret making? This code should capture both the decision itself, the evaluation of its outcome, and the reasons for that evaluation if given. |
| Long-term Benefits* | Beneficial outcomes for the organization or the community that are anticipated to persist after COVID-19. |

**Supplemental Appendix 3.** Interviewee organizational characteristics

| **ID** | **Sector** | **Organization Type** | **Role in Organization** | **Region of NC** | **County Size** | **County Rurality** | **Size of Community/ Constituents** | **Race/Ethnicity of Community/ Constituents** | **Date of Interview** |
| --- | --- | --- | --- | --- | --- | --- | --- | --- | --- |
| G1 | Government | County Management | Assistant County Manager | Western | Counties in metro areas of 250,000 to 1 million population | Metropolitan | 150,000 - 200,000 citizens | Majority White, Minority Black | Nov 9 |
| G2 | Government | County Social Services | Director | Western | Completely rural or less than 2,500 urban population, not adjacent to a metro area | Non-metropolitan | 5000 - 10,000 citizens | Majority White | Nov 4 |
| G3 | Government | County Management | County Manager/ Attorney | Eastern | Urban population of 20,000 or more, adjacent to a metro area | Non-metropolitan | 25,000 - 50,000 citizens | Majority White | Feb 10 |
| G4 | Government | County Management | County Manager | Piedmont | Urban population of 20,000 or more, adjacent to a metro area | Non-metropolitan | 50,000 - 100,000 citizens | Majority White, Minority Black and Hispanic | Feb 17 |
| R1 | Religious | Church | Senior Pastor | Eastern | Counties in metro areas of fewer than 250,000 population | Metropolitan | Thousands of families from ten North Carolina Counties and six Virginia counties. | Majority Black | Sept 14 |
| R2 | Religious | Church | Senior Pastor | Piedmont | Counties in metro areas of 250,000 to 1 million population | Metropolitan | ~10,000 active members | Minority Black and Minority White | Sept 29 |
| R3 | Religious | Church | Senior Pastor | Piedmont | Counties in metro areas of 1 million population or more | Metropolitan | active member count unknown | Majority White | Sept 22 |
| R4 | Religious | Church | Pastor | Piedmont | Counties in metro areas of 250,000 to 1 million population | Metropolitan | active member count unknown | Majority Hispanic | Nov 13 |
| R5 | Religious | Church | COVID-19 Task Force Lead | Piedmont | Counties in metro areas of 250,000 to 1 million population | Metropolitan | ~10,000 active members | Minority Black and Minority White | Oct 7 |
| R6 | Religious | Church | Pastor | Piedmont | Counties in metro areas of 1 million population or more | Metropolitan | active member count unknown | Majority Black | Nov 11 |
| E1 | School | University | Senior Vice Provost for Enrollment Management and Services | Piedmont | Counties in metro areas of 1 million population or more | Metropolitan | ~30,000 total students | Majority White, Minority Black and Asian | Oct 1 |
| E2 | School | Elementary School | Principal | Piedmont | Counties in metro areas of 1 million population or more | Metropolitan | ~500 students | Majority White | Nov 12 |
| E3 | School | Elementary and Middle School | Head of School | Piedmont | Counties in metro areas of 1 million population or more | Metropolitan | 100-200 students | Unknown | Dec 10 |
| E4 | School | University | President | Eastern | Counties in metro areas of fewer than 250,000 population | Metropolitan | ~5,000 total students | Minority White and Black | Oct 14 |
| E5 | School | County School Board | Member | Piedmont | Counties in metro areas of 250,000 to 1 million population | Metropolitan | ~7,000 total students | Majority White, Minority Black or Latino | Oct 28 |
| E6 | School | Count School Board | Superintendent | Eastern | Counties in metro areas of fewer than 250,000 population | Metropolitan | ~25,000 students | Minority White, Black, and Asian | Nov 4 |
| E7 | School | County School Board | Member | Piedmont | Counties in metro areas of 1 million population or more | Metropolitan | ~150,000 students | Majority White, Minority Black and Latino | Nov 6 |
| PH1 | Public Health | County Health Department | Director | Piedmont | Counties in metro areas of 250,000 to 1 million population | Metropolitan | ~300,000 citizens | Majority White, Minority Black | Sept 16 |
| PH2 | Public Health | County Health Department | Director | Western | Counties in metro areas of 250,000 to 1 million population | Metropolitan | 150,000 - 200,000 citizens | Majority White, Minority Black | Sept 9 |
| PH3 | Public Health | County Health Department | Director | Piedmont | Counties in metro areas of 250,000 to 1 million population | Metropolitan | ~300,000 citizens | Majority White, Minority Black or Latino | Sept 25 |
| PH4 | Public Health | County Health Department | Director | Piedmont | Counties in metro areas of 250,000 to 1 million population | Metropolitan | 100,000 - 150,000 citizens | Majority White, Minority Black or Latino | Sept 21 |
| PH5 | Public Health | County Health Department | Director | Piedmont | Counties in metro areas of 1 million population or more | Metropolitan | >300,000 citizens | Majority White, Minority Black or Latino | Sept 21 |
| H1 | Healthcare | Healthcare Society | Director | State-wide | State-wide | State-wide | State-Wide | State-wide | Sept 8 |
| H2 | Healthcare | Statewide Healthcare Association | President | State-wide | State-wide | State-wide | State-Wide | State-wide Association | Sept 11 |
| H3 | Healthcare | Private Health System | Systems Engineer | Eastern | Eastern Region | Multi | ~1,500 bed health system serving ~1M people in Eastern North Carolina | Majority White, Minority Black and Asian | Sept 18 |
| H4 | Healthcare | Healthcare Society | Senior Vice President | State-wide | State-wide | State-wide | State-Wide | State-Wide | Nov 4 |
| H5 | Healthcare | University | Director and Medical Director of Student Health Services | Piedmont | Counties in metro areas of 1 million population or more | Metropolitan | ~30,000 total students | Majority White, Minority Black and Asian | Sept 25 |
| NP3 | Non-profit organization | Community Organization | Vice President of Enterprise Risk Management | Piedmont | Counties in metro areas of 1 million population or more | Metropolitan | 20+ facilities across 5+ counties | Unknown | Oct 29 |
| NP2 | Non-profit organization | Community Organization | Vice President of Operations & Programs | Piedmont, Eastern | Multi-regional | Multi | Thousands of families served each year | Unknown | Dec 10 |
| NP3 | Non-profit organization | Community Organization | Senior Director of Programs | Piedmont | Counties in metro areas of 250,000 to 1 million population | Metropolitan | 100-300 families served each year | Majority Hispanic, Minority White, Asian, and Middle-Eastern | Dec 16 |
| B1 | Business | Real Estate Business | Sales Agent | State-wide | State-wide | State-wide | Unknown | Unknown | Oct 30 |
| B2 | Business | Software Business | Director of Global Public Sector | State-wide | State-wide | State-wide | 10,000 - 15,000 employees | Unknown | Nov 3 |
| B3 | Business | Record Store | Owner | Eastern | Counties in metro areas of fewer than 250,000 population | Metropolitan | Costumers served unknown, located in 10,000 - 15,000 person town | Race/ethnicity of customers served unknown, located in a Majority white town | Jan 21 |
| B4 | Business | Coffee Shop | Owner | Eastern | Counties in metro areas of fewer than 250,000 population | Metropolitan | Costumers served unknown, located in 50,000 - 100,000 person town | Race/ethnicity of customers served unknown, located in a Majority Black, minority White Town | Jan 25 |
| PS1 | Public Safety | County Sheriff Office | Sheriff | Piedmont | Counties in metro areas of 1 million population or more | Metropolitan | ~1M citizens | Majority White, Minority Black | Jan 5 |
| PS2 | Public Safety | County Sheriff Office | Sheriff | Eastern | Counties in metro areas of fewer than 250,000 population | Metropolitan | 150,000 - 200,000 citizens | Majority White, Minority Black | Oct 12 |
| PS3 | Public Safety | County Emergency Services | Director | Piedmont | Counties in metro areas of 250,000 to 1 million population | Metropolitan | >300,000 citizens | Majority White, Minority Black | Nov 4 |
| PS4 | Public Safety | County Emergency Services | Emergency Manager | Piedmont | Counties in metro areas of 250,000 to 1 million population | Metropolitan | >300,000 citizens | Majority White, Minority Black | Nov 10 |
| PS5 | Public Safety | County Emergency Services | Emergency Manager | Western | Counties in metro areas of 250,000 to 1 million population | Metropolitan | 200,000 - 300,000 citizens | Majority White | Feb 10 |
| PS6 | Public Safety | County Emergency Services | Emergency Manager | Eastern | Urban population of 20,000 or more, adjacent to a metro area | Non-metropolitan | 25,000 - 50,000 citizens | Majority White | Feb 11 |
| PS7 | Public Safety | University | Director, Emergency Management and Mission Continuity | Piedmont | Counties in metro areas of 1 million population or more | Metropolitan | ~30,000 total students | Majority White, Minority Black and Asian | Sept 21 |
| T1 | Transportation | City Transportation | Transportation Planner | Western | Counties in metro areas of 250,000 to 1 million population | Metropolitan | 50,000 - 100,000 citizens | Majority White, Minority Black | Jan 7 |
| T2 | Transportation | City Transportation | Bicycle and Pedestrian Coordinator | Piedmont | Counties in metro areas of 250,000 to 1 million population | Metropolitan | 200,000 - 300,000 citizens | Majority White, Minority Black | Jan 15 |
| T3 | Transportation | State Transportation | Traffic Safety Project Engineer | State-wide | State-wide | State-wide | State-wide | State-wide | Jan 5 |

**Supplemental Appendix 4.** Spectrum of key COVID-19-related decisions faced by local North Carolina decision-makers across sectors

| **Sector** | **Decision Category** | | | | | |
| --- | --- | --- | --- | --- | --- | --- |
|  | **Continuing/Suspending In-Person Services** | **Safety Protocols** | **New services/ populations served** | **Testing/Tracing** | **Vaccination** | **Resource Allocation** |
| Business  (N=4) | Retail: Closed shop to public and built online business; Later opened by appointment only  Food: Created outdoor walk-up counter, online ordering and curbside pickup; Later opened inside with clear safety protocols  Retail: Switched to conducting meetings with buyers virtually | Retail/food: Masking, distancing, and sanitizing requirements for customers and staff Real estate: Client self-reporting of COVID exposure; Stopped riding in cars with clients | Retail: Developed social media presence, conducted online fundraisers, and took a political stand  Food: Allowed customers to sponsor care packages to frontline workers  Real estate: Attended more corporate online events | Tech: Implemented system to facilitate contact tracing at scale for clients globally | Tech: Worked with governments and shipping companies on optimization; Modeled public health impacts of different vaccine prioritization schemes | Retail/Food: Bought masks, gloves, hand sanitizer for customers and staff  Tech: Forecasted PPE requirements and used optimization to inform allocation |
| Community Org  (N=3) | Food Distr.: Increased demand, no option to suspend primary services; suspended ancillary services, such as nutrition education  Rec/youth: Cancelled trips, events, conferences, camps, etc. for the first time in org. history | All: Frequently updated safety protocols (related to cleaning, volunteers, travel policies, etc.) in response to changing CDC guidelines; Sought LHD approval | Food Distr.: New delivery methods to accommodate safety protocols and school closings (e.g., individual food boxes delivered by volunteers); CFAP created new partnership structure  Rec/youth: Convened a group of non-profits to support virtual learning/childcare | All: Reported of COVID cases to LHD | -- | All: Shifted nonessential employees to remote work; hired new staff members for phone outreach services |
| County Government  (N=4) | Management: Shut down county completely for 45 days (similar to past hurricane responses)  All: Shifted as many county services as possible remotely | All: Implemented safety protocols for in-person county staff (e.g., social distancing, travel restrictions, glass partitions, expanded office space) | Management: Developed online payment and reservation systems  Social Services: Created a new position to coordinate food delivery to kids on free and reduced lunch | Management: Worked in concert with public health and local hospitals on contact tracing and testing | Management: Providing support to LHD and hospital with staffing and facilities  Social Services: Working with LHD on community engagement | Management: Built partnership to collaboratively source volunteers; Leased new building to accommodate social distancing; Coordinated PPE  Social Services: Redeployed Adult Protective Services to food outreach |
| Healthcare  (N=5) | Associations: Maintained functions remotely  Health system: Championed stay-at-home policy in community; shifted some staff to remote work  Campus Health: Coordinated continuity of care for students leaving campus | Health System: Established symptom screening and PPE protocols for in-person staff | Associations: Connected healthcare organizations with COVID-related information, training for clinician redeployment, and PPE  Health System: Partnered with community orgs to coordinate childcare and address health disparities | Associations: Partnered with CCNC to hire and train additional COVID tracers  Health System: Presented to the state about testing capacity  Campus Health: student quarantine, isolation and testing protocols | -- | Health System: Categorized workers into essential vs. nonessential  Associations: Supplied staff member to state emergency operations to coordinate with health systems; Updated critical care resource allocation protocol; PPE group purchasing |
| Public Health  (N=5) | Informed local school and university reopening/closing decisions; Issued stay-at-home order in advance of the state | Issued mandatory mask order in advanced of the state; Instituted restaurant curfew; Reviewed safety protocols for local org. reopening plans; Developed internal safety protocols for employees | Formed strike teams to deal with homelessness and food insecurity; Hired COVID ambassadors to help businesses understand reopening requirements; Created call center for COVID-related questions; Started community health worker program | Conducted contact tracing and case investigation (part of core LHD function); Some LHDs hosted testing events, while others facilitated testing by health systems and community orgs; Opened temporary homeless shelter for people waiting on test results | Planned safety protocols at mass vaccination clinics; Worked with community partners to build vaccine champions; Partnered with EM to coordinate vaccine logistics | Repurposed staff to COVID tasks; Ensured the well-being and childcare needs of staff were met while working overtime; Purchased equipment for remote work |
| Public Safety  (N=7) | EM: Shut down county completely for 45 days (similar to past hurricane responses)  Law Enforcement: Stopped offering gun permits; Decreased number of arrests | EM: Executed Stay Safe, Stay Home strategy for LTC facilities; Issued warnings for businesses not following protocol  Law Enforcement: Developed staff safety protocols and  detention center quarantine precautions | EM: Set up community centers to address social needs;  Started county fund for small business owners; Created quarantine center for homeless who tested positive; Rented hotel for homeless population  Law Enforcement: Educated homeowners about protective measures due to increases in break-ins | Oversaw logistics for mobile and community testing and contact tracing | Adapted testing model for vaccination; Partnered with EMTs, paramedics, fire, EMS, etc. for administration; Involved in multi-sector stakeholder group to plan rollout and messaging | Trained and redeployed other county employees to emergency services; Updated sick leave and job sharing policies; Ensured PPE access for first responders (partnered with businesses); Forecasted PPE needed to run emergency operation center |
| Religious Org  (N=6) | All: Suspended in-person services for an extended period of time; Several also had schools attached to church, so those were shut down initially as well  About half had resumed in-person services to varying degrees at the time of the interview. | For in-person services: Instituted protocols for social distancing, mask requirements, cleaning/sanitizing; Used ushers to control the flow of people  For staff: Partial remote work, travel restrictions, symptom self-reporting and temperature checks | All: Identified gaps in community social services and worked with other groups to meet those needs; Developed new methods of outreach to church members of all ages; Expanded student tutoring services, health education, and domestic violence response | One church partnered with LHD to hold a drive-through testing event in their parking lot. | -- | Shifted employee roles; Laid off staff for financial reasons; Installed new HVAC systems; Built picnic tables for outdoor gatherings; Added technology for hybrid gatherings and Wifi extenders for the parking lot |
| School  (N=7) | All: Shut down in-person learning (decision made by the state) and transitioned to remote learning; Reopening discussions have been contentious and political;  One county invited certain students back to school who were particularly struggling with online learning. | All: Instituted protocols for social distancing, masks, and symptom reporting; Informed LHD of COVID incidents; COVID coordinators at each school responsible for temperature and symptom checks | Secondary: Provided laptops/technology access/other physical needs to students; Distributed yard signs to stay connected to students  Higher Ed: Developed online course for incoming university students about COVID; Donated PPE and offered free wifi to community members | Higher Ed: Developed testing and isolation procedures for bringing students back | -- | Hired additional cleaning staff; Contentious decisions about remote teaching, keeping staff on, cutting or furloughing jobs, and meeting accommodation requests |
| Transportation (N=3) | All: Transitioned public input sessions to be virtual (Zoom, polling questions, online interactive map); Transitioned nonessential staff to remote work | City: Developed and advertised social distancing and mask requirements on public trails and public transportation; changed buses to be fare free | City: Hired private transportation company to supplement/avoid cutting routes; Lent businesses public space | -- | -- | All: Shifted roles to keep people working and on payroll even if their original job function could not be performed remotely; Hired people to deal with changing operations due to COVID (e.g., more bus drivers) |
